# Longitudinal detection of SARS-CoV-2-specific antibody responses with different serological methods

**DOI:** 10.1101/2021.04.16.21255608

**Authors:** Petra Emmerich, Ronald von Possel, Christoph Josef Hemmer, Carlos Fritzsche, Hilte Geerdes-Fenge, Babett Menge, Claudia Messing, Viola Borchardt-Lohölter, Christina Deschermeier, Katja Steinhagen

## Abstract

Serological testing for anti-SARS-CoV-2 antibodies is used to detect ongoing or past SARS-CoV-2 infections. To study the kinetics of anti-SARS-CoV-2 antibodies and to assess the diagnostic performances of eight serological assays, we used 129 serum samples collected on known days post symptom onset (dpso) from 42 patients with PCR-confirmed COVID-19 and 54 serum samples from healthy blood donors, and children infected with seasonal coronaviruses.

The sera were analyzed for the presence of IgG, IgM and IgA antibodies using indirect immunofluorescence testing (IIFT) based on SARS-CoV-2-infected cells. They were further tested for antibodies against the S1 domain of the SARS-CoV-2 spike protein (IgG, IgA) and against the viral nucleocapsid protein (IgG, IgM) using ELISA.

The assay specificities were 94.4%-100%. The sensitivities varied largely between assays, reflecting their respective purposes. The sensitivities of IgA and IgM assays were highest between 11 and 20 dpso, whereas the sensitivities of IgG assays peaked between 20 and 60 dpso.

IIFT showed highest sensitivities due to the use of the whole SARS-CoV-2 as substrate and provided information whether or not the individual has been infected with SARS-CoV-2. ELISAs provided further information about both the prevalence and concentration of specific antibodies against selected antigens of SARS-CoV-2.

## INTRODUCTION

In the current pandemic, direct pathogen detection via reverse transcription and polymerase chain reaction amplification as well as real-time detection (real-time RT-PCR) is the gold standard for SARS-CoV-2 detection and enables early identification of acute SARS-CoV-2 infections. Serological testing for anti-SARS-CoV-2 antibodies is used to confirm ongoing or past infections with SARS-CoV-2. The detection of antibodies enables confirmation of SARS-CoV-2 infection in patients with typical symptoms and in suspected (asymptomatic) cases. Analysis of anti-SARS-CoV-2 antibodies is typically performed at an advanced stage of infection and thus expands the time frame for COVID-19 diagnostics.

Seroconversion of anti-SARS-CoV-2 antibodies can occur at different points in time after virus contact^1,2^. The features of immune responses to SARS-CoV-2 infections vary significantly between individuals^3^, especially regarding the kinetics, immunoglobulin classes and antigen specificity. In the majority of COVID-19 patients, anti-SARS-CoV-2 antibodies are detectable within two weeks of infection^4–6^. Usually, specific IgM and IgA antibodies are detectable earlier than specific IgG antibodies^5,7,8^. In individual cases, anti-SARS-CoV-2 antibodies are either only detectable more than four weeks after onset of symptoms or not at all due to generally absent antibody secretion^8–10^.

Anti-SARS-CoV-2 antibodies target different structural proteins of SARS-CoV-2. The main immunogens are the spike and nucleocapsid proteins. The highly immunogenic S1 domain of the spike protein of SARS-CoV-2 is a major target for neutralizing antibodies and is being used as the antigen in many serological assays^11^. The immunologically relevant receptor binding domain (RBD) represents another important target antigen for virus-neutralizing antibodies^12^. The nucleocapsid protein (NCP) of SARS-CoV-2 is the antigen with the strongest immune dominance among *Coronaviridae*^*13*^ and contains diagnostically relevant epitopes of SARS-CoV-2. Previous studies suggested heterogeneous binding antibody responses to S1/RBD and NCP viral antigens^14^, and hence the presence of antibodies against one protein of SARS-CoV-2 does not necessarily coincide with the presence of antibodies against another.

Current research is determined to illuminate kinetics of the humoral immune response against SARS-CoV-2, potentially providing guidance on when to use serological tests effectively for screening or monitoring of the infection. Results of serological tests can provide answers to important epidemiological, clinical and virological questions concerning SARS-CoV-2, for instance, on the traceability of infection chains and the role of asymptomatic or presymptomatic transmission. Moreover, exact determination of the course of concentration of IgG antibodies against SARS-CoV-2 before and after vaccination can provide valuable information on the effectiveness of vaccination.

Currently, knowledge about SARS-CoV-2 antibody persistence is scarce, although it would help to understand the possible role of humoral immunity in the protection against reinfection. The aim of this research was to study the kinetics of antibodies against SARS-CoV-2 and to explore the characteristic features of eight serological assays.

## METHODS

### Human serum samples

Panel A comprised 82 sequential and single serum samples from 25 German patients (Table 1). Infection with SARS-CoV-2 was confirmed by PCR^15^ by regional health authorities. These patients had mild to moderate COVID-19 symptoms.

**Table 1:**
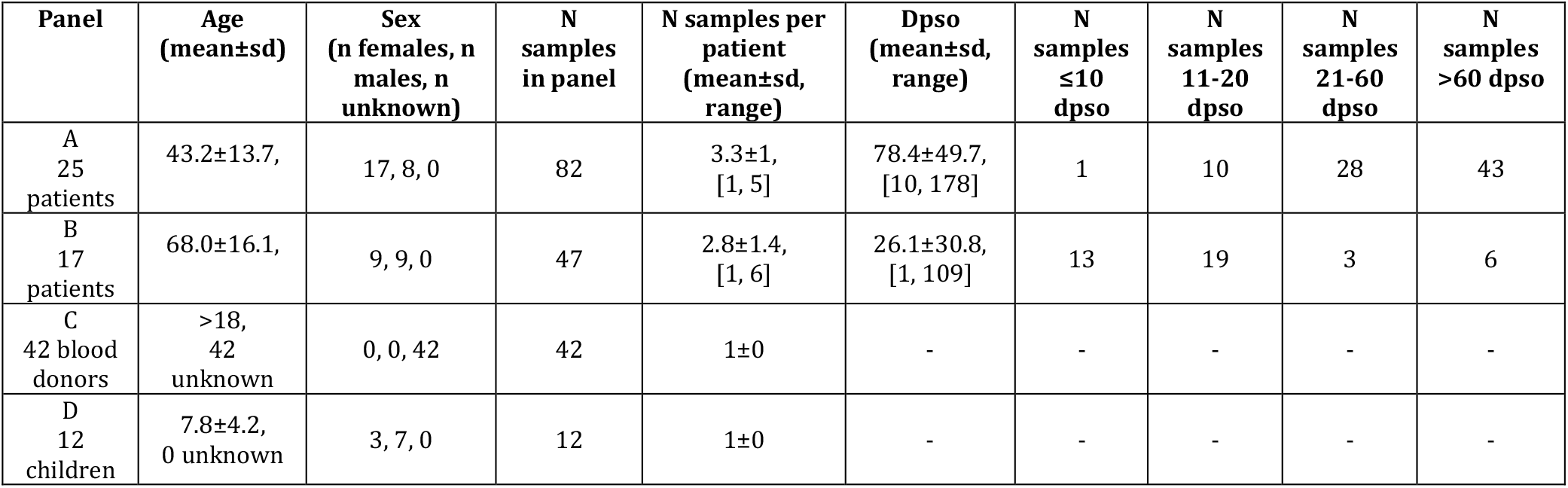
Descriptive information of all panels. Dpso: days after symptom onset. Sd: standard deviation.

Panel B comprised 47 sequential and single serum samples from 17 German patients (Table 1). Infection with SARS-CoV-2 was confirmed by PCR using the Allplex 2019-nCoV Assay (Seegene Inc., Seoul, Korea). These patients required hospitalization.

All patient samples were also serologically precharacterized by IIFT.

Panel C comprised serum samples taken before August 2019 from 42 healthy German blood donors (Table 1).

Panel D comprised serum samples taken between January and March 2020 from twelve German children (Table 1) positive for IgG against seasonal coronaviruses (e.g. HCoV 229-E) by indirect immunofluorescence testing (IIFT, for research use only).

### Detection of anti-SARS-CoV-2 antibodies

The detection of the antibodies against SARS-CoV-2 (genus: *Betacoronavirus*, family: *Coronaviridae*) using IIFT was performed with anti-IgG-, anti-IgA- and anti-IgM-FITC-labelled secondary antibodies on infected Vero E6 cells fixed in acetone-methanol^16,17^. Sample dilutions from 1:20 to 1:80 were screened. In case of a positive result, it was titrated to the final titer. An immunofluorescence signal at titers ≥1:20 was rated as positive and otherwise as negative. Samples were further tested for the presence of antibodies against SARS-CoV-2 with five enzyme-linked immunosorbent assays (ELISA, from EUROIMMUN Medizinische Labordiagnostika AG, Luebeck, Germany). All ELISAs apply viral antigens recombinantly expressed in human cells. The Anti-SARS-CoV-2 ELISA (IgG) and Anti-SARS-CoV-2 ELISA (IgA) are based on the S1 domain of the spike protein of SARS-CoV-2 as antigen, including the immunologically relevant receptor binding domain (RBD), to detect anti-SARS-CoV-2 IgG and IgA antibodies, respectively. The Anti-SARS-CoV-2 QuantiVac ELISA (IgG) was used for quantitative detection of anti-SARS-CoV-2 IgG antibodies by means of a 6-point calibration curve. The Anti-SARS-CoV-2 NCP ELISA (IgG) and Anti-SARS-CoV-2 NCP ELISA (IgM) are based on a modified nucleocapsid protein (NCP) as antigen to detect anti-SARS-CoV-2 IgG and IgM antibodies, respectively. ELISA results were evaluated as recommended by the manufacturer. Borderline results were reported but excluded from the subsequent analyses, since they do not allow secure evaluation and are subjected to retesting by means of other diagnostic methods and/or serological investigation of a follow-up sample in laboratory practice.

The detection of SARS-CoV-2-specific antibody responses was also investigated with respect to the infection phase. Because the diagnostic window for serological testing opens several days after pathogen contact, only samples taken later than ten dpso were considered. The phases were split in 11-20 dpso (early phase of infection; n samples in panel A: 10, panel B: 19), 21-60 dpso (intermediate phase of infection; n samples in panel A: 28, panel B: 3) and >60 dpso (past infection; n samples in panel A: 43, panel B: 6, Table 1).

The overall agreement between qualitative results obtained with the Anti-SARS-CoV-2 ELISA (IgG) and Anti-SARS-CoV-2 QuantiVac ELISA (IgG) was calculated, their degree of agreement was quantified using Cohen’s κ including borderline results^18^, and the statistical association between results was described using Pearson correlation and 95% confidence intervals as determined by Clopper-Pearson interval.

## RESULTS

The sensitivities varied largely between assays (Table 2). The IIFT revealed positive results for anti-SARS-CoV-2 IgG, IgA and IgM antibodies in 94.6%, 72.9% and 65.9% of the patient samples, respectively. The ELISAs detected specific antibodies against S1 IgG and IgA in 75.8% and 80.3% of the patient samples, respectively. Anti-SARS-CoV-2 IgG and IgM antibodies against NCP were detected in 82.0% and 19.8% of the patient samples, respectively. The specificity was 100% by IIFT, Anti-SARS-CoV-2 ELISA (IgG, IgA) and Anti-SARS-CoV-2 NCP ELISA (IgM), while the four remaining assays reached specificities between 92.9% and 97.6%. Cross-reactivities were not observed.

**Table 2:**
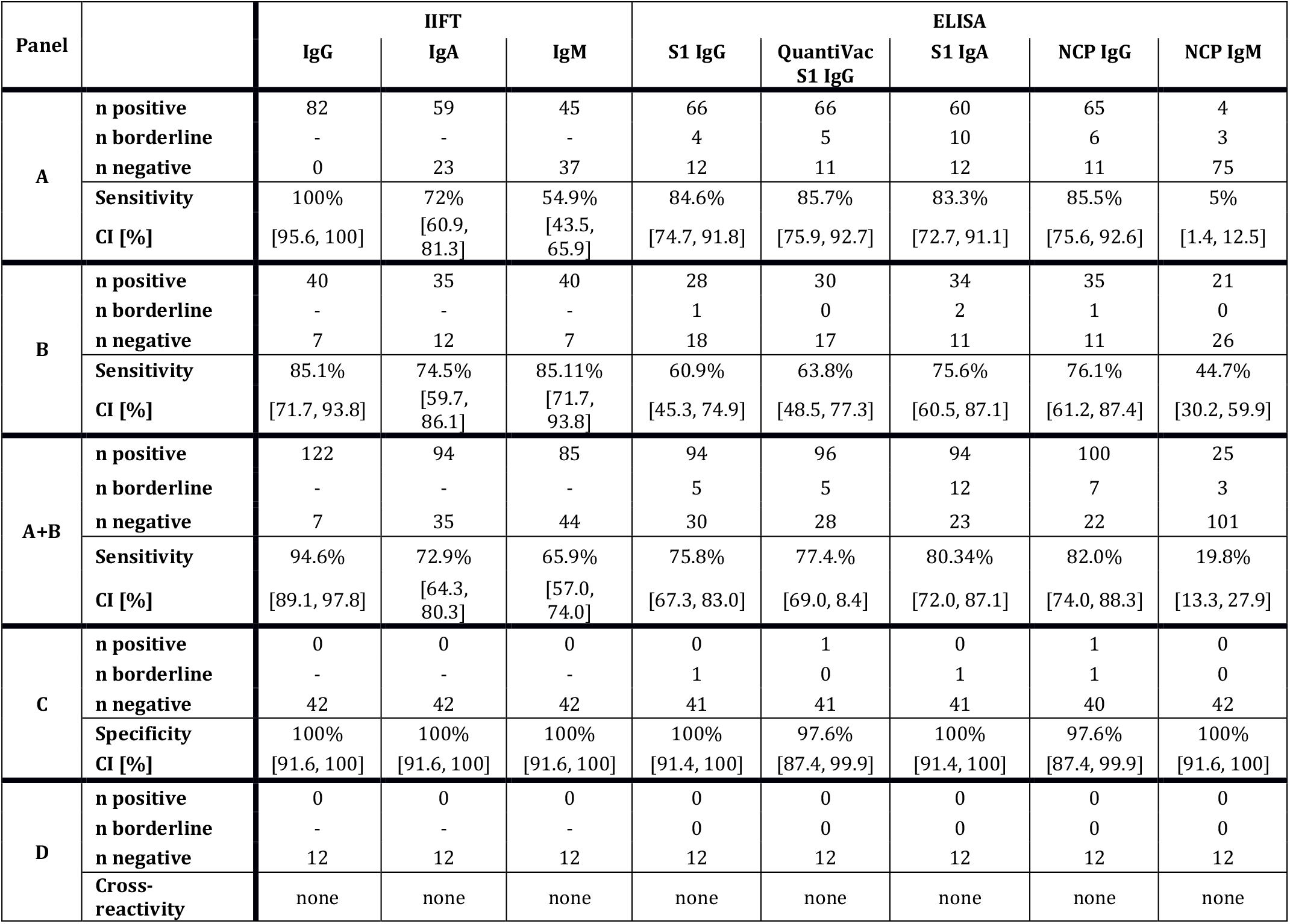
Diagnostic performance of the assays. Sensitivities were determined based on panel A (n samples = 82), panels B (n samples = 47) and panels A+B (n samples = 129). Specificities were determined based on panel C (n samples = 42). Cross-reactivities were determined based on panel D (n samples = 12). CI: 95% confidence interval. Borderline ELISA results were excluded for calculation of the sensitivity and specificity.

The sensitivities of IgA and IgM assays were highest in the early phase of infection, while positive results for IgG antibodies occurred most often in the intermediate phase (Table 3, Figure 1).

**Table 3:** Number of positive results and sensitivity (%) per infection phase based on 109 serum samples from panels A+B taken later than ten days after onset of symptoms (dpso). For ELISA, the number of borderline results are reported in brackets but were excluded for calculation of the sensitivity.

| Phase<br>[dpso] | N<br>samples | IIFT |  |  |  |  |  | ELISA |  |  |  |  |  |  |  |  |  |
| --- | --- | --- | --- | --- | --- | --- | --- | --- | --- | --- | --- | --- | --- | --- | --- | --- | --- |
|  |  | IgG |  | IgA |  | IgM |  | S1 IgG |  | QuantiVac<br>S1 IgG |  | S1 IgA |  | NCP IgG |  | NCP IgM |  |
|  |  | n | % | n | % | n | % | n | % | n | % | n | % | n | % | n | % |
| 11-20 | 29 | 28 | 96.6 | 27 | 93.1 | 28 | 96.6 | 19<br>(2) | 70.4 | 20<br>(1) | 71.4 | 24<br>(2) | 88.9 | 25<br>(0) | 86.2 | 14<br>(1) | 50.0 |
| 21-60 | 31 | 31 | 100 | 31 | 100 | 27 | 87.1 | 28<br>(1) | 93.3 | 28<br>(1) | 93.3 | 24<br>(2) | 82.8 | 30<br>(0) | 96.8 | 4<br>(0) | 12.9 |
| >60 | 49 | 48 | 98.0 | 22 | 44.9 | 15 | 30.6 | 40<br>(2) | 85.1 | 40<br>(3) | 87.0 | 33<br>(8) | 80.5 | 35<br>(6) | 81.4 | 0<br>(2) | 0.0 |
| ≥11 | 109 | 107 | 98.2 | 80 | 73.4 | 70 | 64.2 | 87<br>(5) | 83.7 | 88<br>(5) | 84.6 | 81<br>(12) | 83.5 | 90<br>(6) | 87.4 | 18<br>(3) | 17.0 |

**Figure 1:**
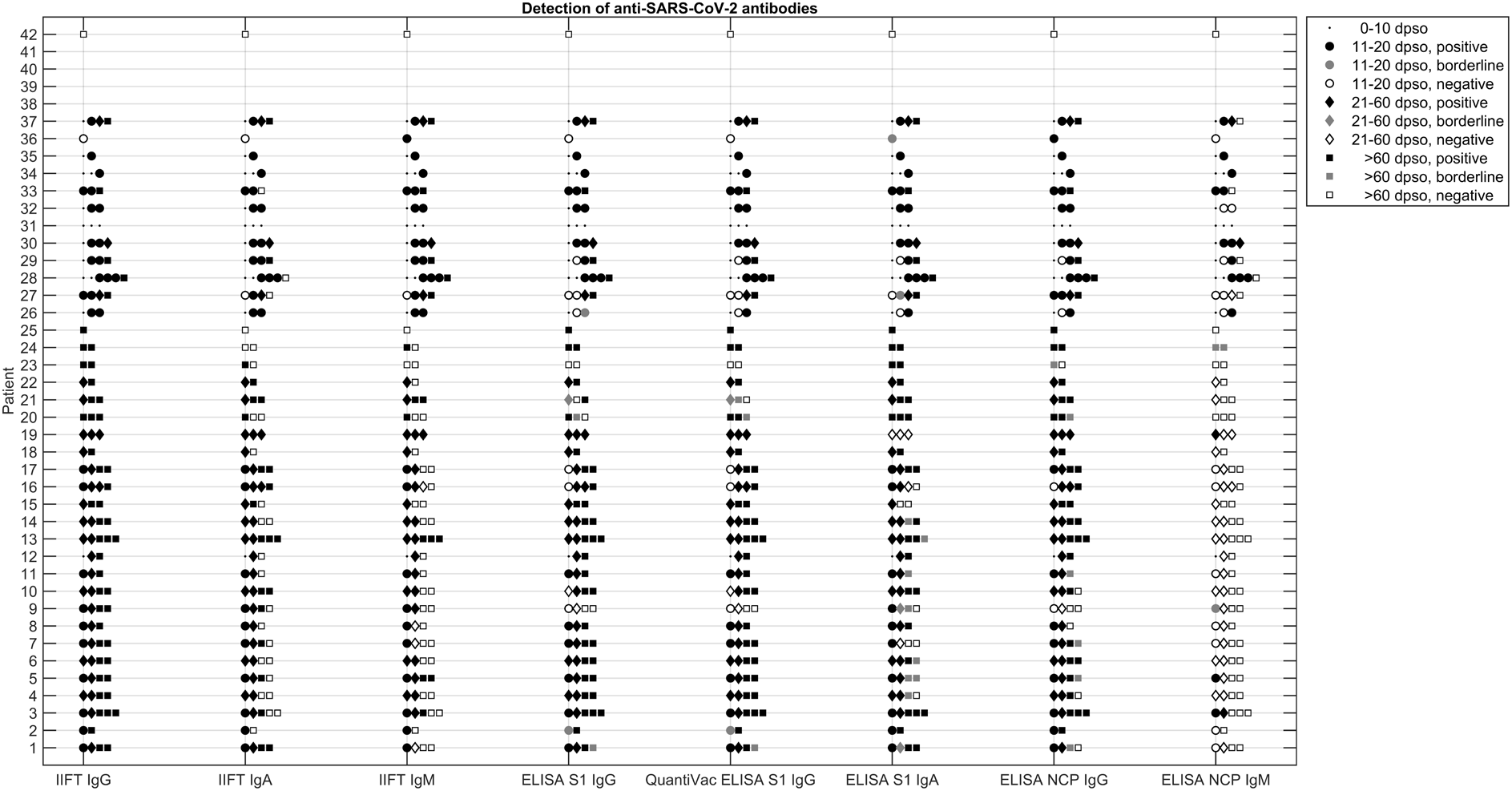
Longitudinal detection of SARS-CoV-2-specific antibody responses in serum samples from panel A (25 patients, 82 samples) and B (17 patients, 47 samples, results of six samples with unknown dpso are not displayed) with respect to phase of infection. Dpso: days after onset of symptoms.

Positive results for anti-SARS-CoV-2 IgG antibodies against S1 reached a peak during the intermediate phase of infection. In contrast, positive results for anti-SARS-CoV-2 IgA antibodies as measured by IIFT showed an initial peak followed by a pronounced decrease after 60 dpso. During the course of infection, the number of positive results for anti-SARS-CoV-2 IgM antibodies dropped as measured both by ELISA and IIFT.

In the early phase of infection (11-20 dpso), IgG and IgA antibodies against S1 of SARS-CoV-2 were detected in 70.4% and 88.9% of the samples (n=29), respectively, while IgG and IgM antibodies against NCP were detected in 86.2% and 50%, respectively. The IIFT detected SARS-CoV-2 specific IgG, IgA and IgM antibodies in 96.6%, 93.1% and 96.6% of the samples, respectively.

In the intermediate phase of infection (21-60 dpso), IgG and IgA antibodies against S1 of SARS-CoV-2 were detected in 93.3% and 82.8% of the samples (n=31), respectively, while IgG and IgM antibodies against NCP were detected in 96.8% and 12.9%, respectively. The IIFT detected specific IgG, IgM and IgA antibodies in 100%, 87.1% and 100%, respectively.

In the late phase of infection (>60 dpso), IgG and IgA antibodies against S1 of SARS-CoV-2 were detected in 85.1% and 80.5% of the samples (n=49), respectively, while IgG and IgM antibodies against NCP were detected in 81.4% and 0%, respectively. The IIFT detected specific IgG, IgA and IgM antibodies in 98%, 44.9% and 30.6%, respectively.

Overall, in samples taken later than 10 dpso, IgG and IgA antibodies against S1 of SARS-CoV-2 were detected in 83.7% and 83.5% of the samples (n=109), respectively, while IgG and IgM antibodies against NCP were detected in 87.4% and 17%, respectively. The IIFT detected specific IgG, IgA and IgM antibodies in 98.2%, 73.4% and 64.2%, respectively.

The Anti-SARS-CoV-2 QuantiVac ELISA (IgG) showed a high total agreement (98.9%, Table 4), an almost perfect degree of agreement (κ=0.93, 95% confidence interval: [0.87, 0.98]) of qualitative results with the Anti-SARS-CoV-2 ELISA (IgG) as well as a high correlation (r=0.98, p<0.001) between (semi)quantitative results (Figure 2).

**Table 4:**
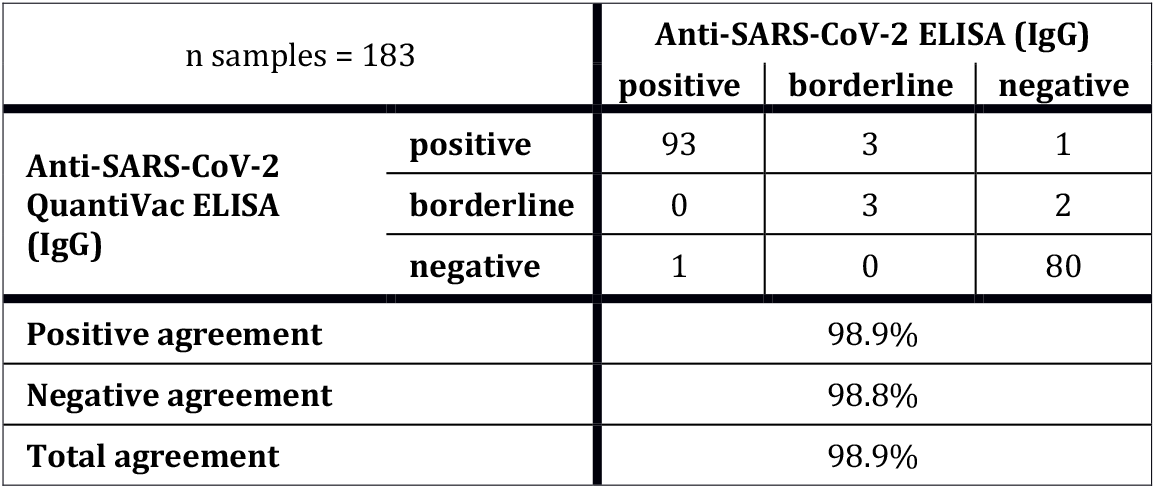
Agreement between qualitative results of Anti-SARS-CoV-2 ELISA (IgG) and Anti-SARS-CoV-2 QuantiVac ELISA (IgG) based on 183 serum samples (panels A, B, C and D).

**Figure 2:**
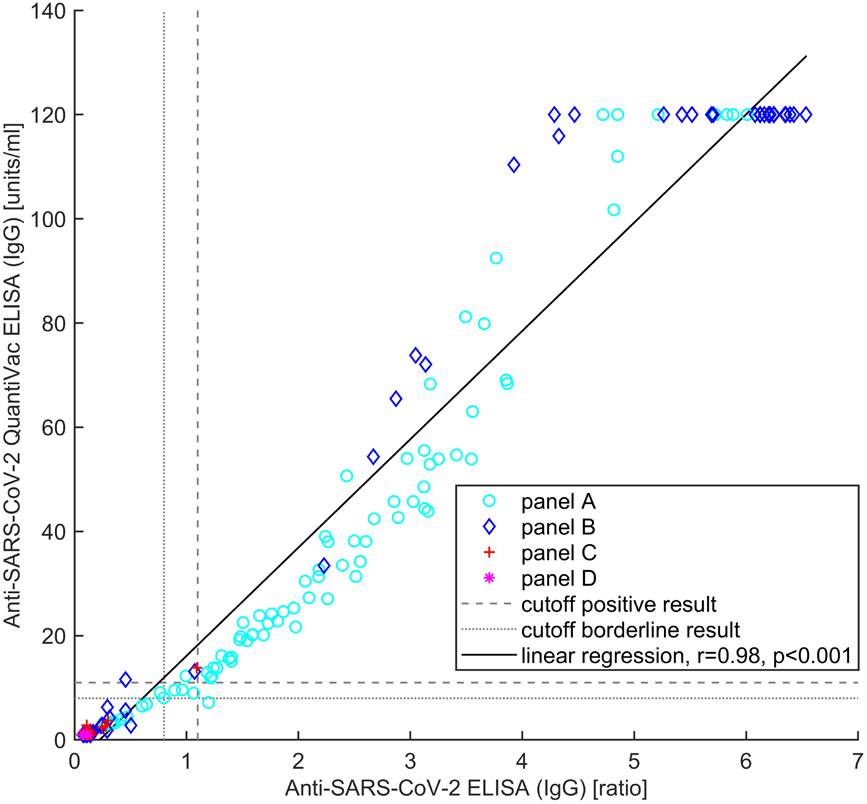
Correlation between semiquantitative results of Anti-SARS-CoV-2 ELISA (IgG) and Anti-SARS-CoV-2 QuantiVac ELISA (IgG).

## DISCUSSION

The aim of this research was to study long-term kinetics of antibodies against SARS-CoV-2 and to assess the characteristic features of different serological methods. We present findings of the temporal profiles of IgG, IgA and IgM antibody responses against SARS-CoV-2 determined in sera from patients with mild to severe COVID-19 by means of eight serological assays.

The sensitivities varied between assays and phases of infection but corroborated that the different substrates used in the assays serve different purposes. Due to the use of the whole SARS-CoV-2 as substrate and the precharacterization by IIFT, the IgG IIFT showed, overall, the highest sensitivity (94.6%) when testing all patient samples, independent of the infection phase (Table 1).

The ELISAs, in contrast, provide information about the prevalence of specific antibodies against selected antigens of SARS-CoV-2. Hence, lower sensitivities of the S1-specific ELISAs compared to the NCP-specific ELISA probably reflect the known fact that not all infected individuals produce antibodies against the S1 domain of SARS-CoV-2^10^. Importantly, previous research showed that responses of specific IgG against S1 and NCP may be heterogeneous between individuals, time-delayed and do not always coincide with each other^8,12,14^. In the present panels, the prevalence of specific IgG antibodies against NCP in the early phase of infection was higher than that against S1 (Table 2). However, the findings of the current study do not support previous research by Herroelen et al who undertook a comparative evaluation of commercial SARS-CoV-2 serological assays and observed no clear differences in the seroconversion kinetics of antibodies targeting SARS-CoV-2 S and N protein epitopes between severe and milder SARS-CoV-2 infections.

Exclusively in the early phase of infection, the prevalence of specific IgA antibodies against S1 was higher than that of specific IgG antibodies against NCP as well as S1. This observation reflects that of Okba et al^1^. However, it is contrary to a previous study that showed a higher sensitivity of the Anti-SARS-CoV-2 IgG compared to the Anti-SARS-CoV-2 IgA ELISA in patient samples taken later than fourteen dpso^19^, whereby the discrepancy might be due to heterogeneous definitions regarding the early phase of infection.

The IgA IIFT showed a pronounced decrease in the antibody detection rate after 60 dpso, which was not observed for the IgA ELISA (Table 2). A possible explanation for this might be that the IgA antibody response against the S1 protein largely remains constant, while the production of IgA antibodies against other antigens of SARS-CoV-2 decreases.

More patients were seropositive for IgM by IIFT than by ELISA (Table 1, Table 2), which could be accounted for by a low sensitivity of the NCP IgM ELISA, warranting further investigations. However, the continuously low sensitivity of the NCP-specific IgM ELISA (Table 2) is in accordance with previous results indicating a sensitivity of 55% at week three to four after disease onset^20^. Liu et al also observed a higher sensitivity of an ELISA based on the spike protein compared to an NCP-based ELISA for detection of IgM antibodies^8^. Two months after symptom onset, we observed a decline in the sensitivity of both IgM-specific assays (Table 2). Independent of the serological method, the two IgM-specific assays reached maximal sensitivities between 11-20 dpso (Table 2) and could therefore especially be applied to detect antibodies in samples taken during the early phase of infection. If patients develop specific IgM against NCP, these antibodies seem to be present for only a short time during the early phase of infection. A sharp decline in the IgM prevalence is to be expected because isotype switching of virus-specific B-cells from IgM to IgG antibody production causes a decline in circulating IgM^21^. The facts that SARS-CoV-2-specific IgM is detected mostly in the early infection phase but only in rare cases^22^ invites the question whether all isotypes should be measured during serodiagnostics.

The agreement analysis revealed a very high correlation between results obtained with the Anti-SARS-CoV-2 ELISA (IgG) and the QuantiVac ELISA (IgG). The two samples that showed inconsistent qualitative results between these assays (Table 3, Figure 2) were taken relatively early and late (7 and 116 dpso) in the course of the disease. An explanation for these inconsistencies might be that the assays were incubated using the same aliquot but on different days, hence the experimental conditions might have differed slightly. Another reason might be that the artificial division between positive and negative results does not match the natural range of activity of some samples.

In general, the use of cells infected with the whole SARS-CoV-2 as substrate has the great advantage of obtaining a high sensitivity due to presence of the complete antigenic spectrum, as evident in the present IIFT results. This is, however, linked to the disadvantage that a positive IIFT result does not allow for a conclusion on the molecular identity of the antigen(s) binding the antibody. In contrast, recombinant cell substrates used in the ELISA technique are ideally suited for the detection and precise identification of antibodies against selected proteins of SARS-CoV-2 such as S1/RBD and NCP. During the purification required for Anti-SARS-CoV-2 NCP ELISA production, tertiary or quaternary structured epitopes are often destroyed or weakened. Nevertheless, a selective loss of reactivity does have advantages, since undesired antibody binding aside from the recombinant target protein can be suppressed. Thereby, the specificity of the ELISA can be improved, which was evident in the present results. Moreover, the ELISA technique has the advantage of yielding results in numeral form, which allows an objective evaluation of results. The use of SARS-CoV-2 IIFT is (currently) reserved for specialized research laboratories with high biosafety restrictions due to the handling of the full virus, and, compared to other serological techniques, IIFT is less implemented in standard diagnostic laboratories.

The presence of anti-SARS-CoV-2 S1/RBD IgG antibodies seems to correlate with the development of both virus neutralization and immunity^1,3,23^. Previous research found that titers of neutralizing antibodies were significantly correlated with the levels of anti-RBD IgG^12^, and RBD-specific IgG titers were suggested as a surrogate of neutralization potency against SARS-CoV-2 infection^24^. Nevertheless, it is possible that a patient does not develop antibodies against S1 of SARS-CoV-2, but only against NCP. However, this would suggest that neutralizing antibodies might not be present since binding antibodies against NCP seem to correlate to a lesser degree with immunity than binding antibodies against S1/RBD^25^. The development of immunity to SARS-CoV-2 is induced both by the humoral and the cellular immune response, whereby especially IgG directed against the S1 subunit of the SARS-CoV-2 spike protein and specific long-lived T cells are of great interest, as they are suspected to play the most relevant roles in virus neutralization and sustained immunity. A combination of serological tests to quantify both the interferon-gamma release by SARS-CoV-2-specific T cells, stimulated by SARS-CoV-2 specific antigens, and the presence of anti-S1/RBD IgG antibodies will enable differentiated investigation of the immune response in the progression of infection and vaccination. Especially the determination of relevant antibody concentrations will probably be one of the most important instruments for determining the vaccination success, although it is yet unknown how many antibodies against S1/RBD an individual must produce after vaccination to be protected from COVID-19. Surrogate neutralization assays detect circulating neutralizing antibodies against SARS-CoV-2 that block the interaction between the RBD of SARS-CoV-2 with the ACE2-cell surface receptor of the human host cell, thus supporting a quick diagnostic statement about the degree of immunity. In contrast to plaque-reduction neutralization tests, which require handling of the virus, surrogate neutralization assays can easily be integrated in the laboratory routine and do not require biosafety level 3 laboratories.

A detailed analysis of potential associations between antibody kinetics and disease severity was not performed because symptoms were not systematically recorded and the disease severity could therefore not be rated other than that patients in panel A had no or mild symptoms and patients in panel B required hospitalization. Nevertheless, the assay sensitivities were reported also for each panel separately (Table 1). Analysis of temporal profiles was performed on samples from both patient panels because the distribution of samples in the three infection phases was unbalanced between panels A and B (Table 1).

ELISA or immunoblot techniques might be used in future to differentiate between reactivities against distinctive SARS-CoV-2 antigens, which might be useful for determination of biomarkers indicative of early or late infection phases.

In summary, evidence of this study emphasizes that the assays have different advantages as well as intended purposes. ELISAs provide an insight into the prevalences of specific antibodies against selected antigens of SARS-CoV-2. Due to the heterogeneity of individual antibody responses, ELISA may not yield positive results for all patients but a combination of ELISAs with different antigens can reduce this diagnostic gap. The three Anti-SARS-CoV-2 ELISAs that detect IgG antibodies can be used to confirm pathogen contact, starting from week two of the infection, to monitor the humoral response following an acute infection confirmed by direct detection and to detect past infections. The highly immunogenic S1 domain of the spike protein of SARS-CoV-2 is a major target for neutralizing antibodies and showed good correlation with different test systems for the detection of neutralizing antibodies^19,26,27^. IgA-specific ELISAs might further be used to monitor the immune response in COVID-19 patients. IIFT showed highest sensitivities due to the use of the whole SARS-CoV-2 as substrate and provide information whether or not an individual has been infected with SARS-CoV-2.

## Data Availability

The data that support the findings of this study are available from the corresponding author, upon reasonable request.

## Ethics Statement

The observational study has been performed in agreement with the Declaration of Helsinki. It has been approved by the Ethics Review Board of the University Medicine Rostock (registration number: A2020-0086) and the Ethics Review Board of the Medical Association Hamburg (registration number: 2020-10162-BO-ff).

The samples from healthy adults (panel C, Table 1) were collected via blood donation. Diagnostic leftover samples after completion of all diagnostic measures from children (panel D, Table 1) were collected by a routine clinical laboratory (Lübeck, Germany). All samples were processed anonymously.

## Acknowledgements

The authors thank Neele Pekarek of BNITM for supporting performance of the analyses at the facilities of the BNITM.

The authors thank Marieke Schulz of EUROIMMUN Medizinische Labordiagnostika AG for supporting laboratory analysis, which was funded by EUROIMMUN Medizinische Labordiagnostika AG, Luebeck, Germany, in accordance with Good Publication Practice (GPP3) guidelines (http://www.ismpp.org/gpp3).

All probands and patients are gratefully acknowledged for providing samples and participating in this study.

## Author contributions

Petra Emmerich: Conceptualization, Investigation, Funding Acquisition, Project Administration,

Writing -Review & Editing

Ronald von Possel: Investigation, Review & Editing

Christoph Hemmer: Investigation, Data curation, Review & Editing

Hilte Geerdes-Fenge: Investigation, Data curation, Review & Editing

Carlos Fritzsche: Investigation, Data curation, Review & editing

Babett Menge: Investigation, Data curation, Writing -Review & Editing

Claudia Messing: Resources, Writing – Review & Editing

Viola Borchardt-Lohölter: Formal analysis, Visualization, Writing – Original Draft Preparation Christina Deschermeier: Funding Acquisition, Project Administration, Review & Editing

Katja Steinhagen: Resources, Project Administration, Writing – Review & Editing

